# A feasibility study of BEhavioural ACtivation for Haemodialysis: The BEACH Study protocol

**DOI:** 10.64898/2026.06.25.26356529

**Authors:** C. Carswell, S. Metcalfe, R. Agyekum, F. Awan, S. Bhandari, K. Bramham, J. Chilcot, J. Millar, L. Gega

**Affiliations:** Department of Health Sciences, University of York, York, UK; Tees, Esk and Wear Valley NHS Foundation Trust, York, UK; King’s College Hospital, London, UK; Patient and Public Involvement Co-investigator; Hull University Teaching Hospitals NHS Trust; King’s College London, London, UK; Hull York Medical School

**Keywords:** Haemodialysis, depression, psychological intervention, feasibility study

## Abstract

**Introduction:** People with kidney failure receiving haemodialysis experience significantly high rates of depression. However, there is a lack of evidence on how to treat depression in this population. A significant barrier to effective treatment is the high treatment burden of haemodialysis, which makes additional appointments prohibitive. Behavioural activation (BA) is an evidence-based brief therapy for depression that has been delivered in a variety of different clinical settings. However, it has not previously been evaluated in the haemodialysis setting.

**Methods:** This study aims to evaluate the feasibility and acceptability of a cluster randomised controlled trial (cRCT) of intradialytic BA for people with kidney failure. The study consists of three main components: A pilot cRCT, where we will recruit 52 people who are receiving haemodialysis and experiencing symptoms of depression across two sites. Patients will be cluster-randomised to either BA or usual care and will be followed up for three months; a qualitative process evaluation using semi-structured interviews to explore the experiences of patients, healthcare professionals and carers; and a feasibility economic evaluation exploring the feasibility of collecting healthcare resource use data.

**Results:** Key findings will include the feasibility of screening and recruiting participants, participant retention, completion of clinical outcome measures, and the acceptability of the intervention.

**Conclusion:** If feasible, the next step will be to conduct a definitive, adequately powered cRCT to determine the effectiveness of the intervention in this population, so that we can improve the identification and management of depression for people with kidney failure.

## Introduction

Depression has serious implications for the overall health of people receiving haemodialysis (1–3). Depression is associated with decreased quality of life and increased morbidity (4) and is an independent predictor of mortality, in part due to the impact that depression has on adherence to treatment (4). People experiencing depression are less likely to be adherent to their treatment regimen(4), increasing their risk of hospitalisation, morbidity and death (5). Therefore, the issue of depression must be addressed amongst people receiving haemodialysis, to improve long-term outcomes for these patients, their families and carers and reduce the economic burden on the health service.

Improving mental health support for people receiving haemodialysis has been identified as a priority by Kidney Care UK’s Psychosocial Health – A Manifesto for Action (6). Currently, there is limited evidence on the effectiveness of antidepressant medication amongst people receiving haemodialysis (7), in part because of the difficulties in trial design and the adverse events associated with medication (3, 7–9).

Cognitive behavioural therapy (CBT), another evidence-based treatment for depression, appears to be more effective at reducing symptoms of depression among people with kidney disease (10, 11). However, issues surrounding long-term implementation in practice, such as the need for a trained clinical psychologist to deliver the intervention (11), the requirement to attend separate appointments (either in-person, online, or over the phone) (12, 13), raise concerns about feasibility when resources are significantly limited, and patients already face a substantial treatment burden (7, 8).

Behavioural activation (BA) is an evidence-based psychological therapy that forms a key part of CBT. BA is based on making small changes in daily life to disrupt negative cycles of thoughts, feelings, and behaviour (13). While there is evidence that CBT can improve depression among people with chronic kidney disease (CKD), treatment burden limits the accessibility of CBT and there is a lack of evidence supporting the effectiveness of BA alone (10); however, it has been shown to be effective in the treatment and prevention of depression in the general population (14, 15), and can be more cost-effective than traditional CBT (16). Consequently, BA is recommended by the National Institute for Health and Care Excellence (NICE) guidelines on the management and recognition of depression in adults with chronic physical health problems (17).

There is a growing recognition of the need to integrate mental and physical health services, particularly to benefit people who have long-term physical health conditions, such as kidney failure (3, 18–20). This integration is of particular importance for people receiving haemodialysis, due to the impact of the treatment on mental health (21, 22). A specific benefit of BA for this population is that it can be delivered outside of mental health services and has been successfully integrated into other services, such as primary care, where it has been delivered by trained non-specialists (23–25). However, while there is a need to establish the effectiveness of BA for reducing symptoms of depression among people receiving haemodialysis, the feasibility of integrated BA in haemodialysis settings is unknown (10). Therefore, a feasibility study is needed to determine whether the integrated delivery of BA is possible and acceptable before any assessment of effectiveness (26, 27, 29, 30).

### Aims and objectives

This study aims to establish the feasibility and acceptability of a behavioural activation intervention, integrated into haemodialysis care, to address symptoms of depression and anxiety amongst people with kidney failure who are receiving maintenance haemodialysis. To achieve this, we will meet the following objectives:

- Determine the feasibility of recruiting, randomising, and retaining participants, including screening for depression using the PHQ-4, in a definitive cRCT.
- Evaluate the feasibility of data collection methods within a cRCT.
- Evaluate the feasibility of conducting a future economic evaluation of an integrated behavioural activation intervention.
- Explore the acceptability of the integrated behavioural activation intervention for patients, healthcare professionals and informal carers.
- Refine the process for evaluating intervention fidelity in a definitive cRCT.

## Materials and Methods

### Study Design

This study comprises a comprehensive mixed-methods feasibility study with three main components: a feasibility cluster randomised controlled trial (cRCT), a feasibility economic evaluation, and a process evaluation. This protocol is reported in accordance with the SPIRIT checklist for trial protocols (28). This study has been prospectively registered on ISRCTN (ISRCTN74685186) (29), and received ethical approval from the London – Dulwich Research Ethics Committee (REC Reference: 25/LO/0920).

### Research setting

This study will take place in two haemodialysis units in England, one in the North and one in London. Recruiting from two different geographic areas with varying levels of deprivation will enable us to explore different clinical and socioeconomic factors that may influence recruitment and participation in a definitive trial. Recruitment and data collection will take place in the summer and autumn of 2026.

### Feasibility cRCT

#### Study Population

**Inclusion criteria:**

Patients will be eligible for participation if they:

◦ Are over 18 years of age.
◦ Are receiving maintenance in centre haemodialysis for > 3 months and will continue to receive haemodialysis in the unit for the duration of the study.
◦ Have the capacity to provide informed consent.
◦ Score ≥ 3 on the screening PHQ-4

Depression is not regularly screened for within practice, despite recommendations from recent reports on depression in people with kidney disease (6). There is also evidence that people receiving haemodialysis are reluctant to undergo screening for depression (30). Therefore, we will be unable to screen using routinely collected data. Instead, we will screen all potentially eligible patients at each unit using the PHQ-4. This is a valid ultra-brief tool that can detect both anxiety and depression. Patients who score three or higher on the PHQ-4 will be invited to participate in the study (31). All patients who screen positive using this threshold will be provided with information on support services

**Exclusion criteria:**

Patients will be ineligible for participation if they:

- Started antidepressant medication or forms of talking therapy within the previous three months.
- Are receiving haemodialysis for acute kidney injury
- Do not have the capacity to provide informed consent
- Are unable to understand and communicate effectively in English.

#### Sample size

As this is a feasibility study, a power calculation was not conducted, as we are not looking to measure effectiveness. There is no consensus on the appropriate sample size for a feasibility study (32, 33). However, a total sample size of 52 people would allow us to estimate the recruitment rate with a precision of ±9% (assuming a previously observed 40% recruitment rate and a 95% confidence interval). This provides an appropriate level of precision to inform the likely number of people the full-scale trial might need to screen. Given that we have four clusters, this would also allow for a minimum of 12 participants per cluster, with a total of 26 participants recruited from each site (34).

#### Recruitment

Following the initial screen, participants will be approached by the researcher, who will explain the study, provide the participant information sheet, and answer any questions they may have. At the next scheduled haemodialysis appointment, the researcher will approach the patient again and obtain informed consent if they are interested in participating. Screening logs will be used to capture the proportion of patients who are eligible for screening, the proportion of people who are eligible following screening, and the proportion of participants who consent and are randomised.

#### Randomisation

Following the collection of baseline data from all enrolled participants, patients will be grouped into clusters based on their dialysis shift patterns. These shift patterns will be randomly assigned using Stata19 (35) to either the control group, where enrolled participants will receive enhanced usual care, or the intervention group, where enrolled participants will receive the intervention. We will explore the feasibility of cluster randomisation according to the dialysis shift pattern.

Cluster randomisation by dialysis shift pattern will be used to prevent inter-arm contamination, as the intervention is delivered bedside. We will monitor shift changes and potential contamination (e.g., control participants observing intervention delivery) to assess the feasibility of this randomisation strategy.

**Figure 1.**
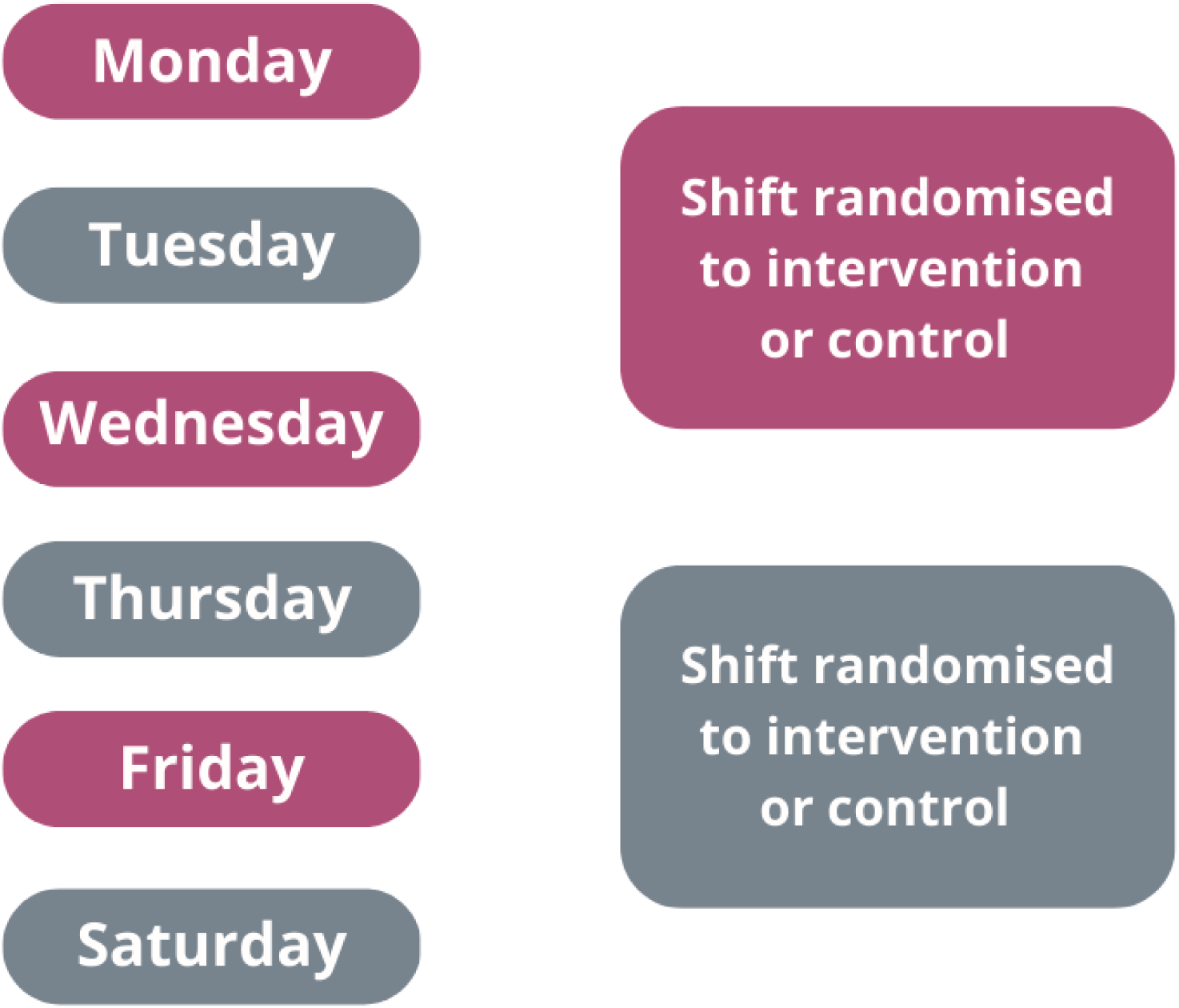
Proposed dialysis shift clusters at each site for randomisation.

#### Intervention

A BA workbook was adapted from the ComBAT intervention (36) and other BA interventions specifically designed for individuals with long-term conditions. This workbook was then shared with our PPI panel for further adaptation to the haemodialysis setting. Each participant allocated to the intervention group will receive at least 6 one-to-one sessions, delivered at their bedside during their haemodialysis treatment. Participants will receive a BA workbook to support the content of each one-to-one session and to refer to between sessions. At the end of each session, the facilitator and participant will plan take-away activities to do outside of the sessions.

**Figure 2.**
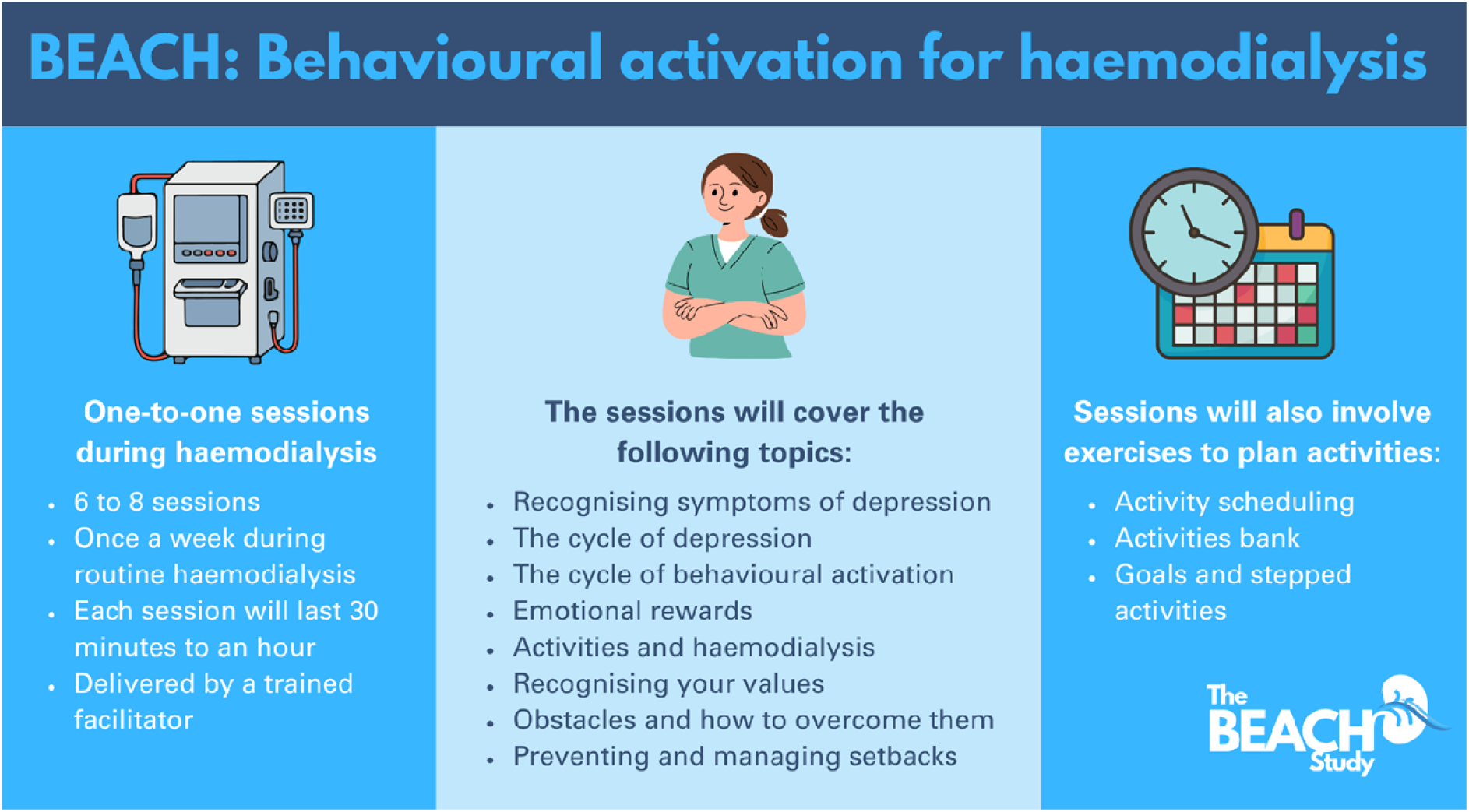
Overview of the BEACH intervention

Using a standardised training programme, we will train kidney health professionals to deliver the BA intervention as facilitators. The training will involve one face-to-face workshop that includes both a theoretical component explaining what BA is and how it works and a skills-based component featuring case scenarios and role-plays. While the content will be organised into 5 key modules, the pacing of session content will be tailored to the participants’ needs. Following the training, the BA facilitators will be offered supervision as needed, both individually and in groups, at most once a week for 30 minutes and at least once a month for 1 hour.

#### Intervention fidelity

Intervention fidelity will be evaluated using session logs. The BA facilitators will be asked to complete a session log following each session, which will record the session length and include a menu of intervention components (e.g., that could be delivered during the session. These will be reviewed to identify whether core aspects of BA have been delivered as intended.

At follow-up, all participants will be invited to complete the Aspects of Care Questionnaire to help assess fidelity and contamination. The questionnaire has eight items that correspond to general activities that may have taken place within both randomisation arms (e.g. “I had an opportunity to talk about my current problems/general life”) as well as BA-specific activities (e.g., “I wrote down things I did for pleasure and necessary tasks on a weekly calendar”). We will compare responses between BA and control group participants to determine whether BA participants received the core components of the intervention as intended (fidelity) and whether control participants inadvertently received BA components (contamination) (36).

#### Usual Care

There is no standardised usual care for depression provided within haemodialysis units across the UK, and few units implement NICE guidelines for the assessment and management of depression in long-term conditions. Usual care typically takes the form of internal or external signposting to services, within the NHS or primary care, or through third-sector providers such as Kidney Care UK (6). Therefore, the control group participants will be provided with information on available services and sign-posted to the relevant service.

#### Data collection

##### Baseline demographic and clinical data

We will collect data on participants’ age, sex, ethnicity, employment status, Index of Multiple Deprivation, time on dialysis, primary kidney diagnosis, and comorbidities. We will also collect self-reported data on whether participants have received a diagnosis of depression at any time in their life, if they are receiving any treatment for mental health conditions, and what that treatment entails.

##### Feasibility measures

The main outcome of the pilot cRCT is feasibility, which will be evaluated by collecting descriptive data on recruitment, retention, and participants’ willingness to be randomised, in accordance with the CONSORT guidelines for feasibility and pilot trials (40). The following data will be collected:

- **Willingness to screen:** The proportion of individuals who agree to undergo the initial screening for depression with the PHQ-4.
- **Eligibility Criteria Suitability:** The number of individuals excluded from the trial and the specific reasons for exclusion will be recorded to assess the appropriateness and practicality of the inclusion and exclusion criteria.
- **Willingness to Participate/ Recruitment rate:** The proportion of eligible individuals who consent to participate in the trial will be recorded as a measure of feasibility and acceptability.
- **Retention Rates:** The proportion of randomised participants who remain in the study and complete the follow-up assessments will be documented. Follow-up contact will be made, where possible, to ascertain the reasons for discontinuation in cases of participant drop-out.
- **Intervention adherence:** The proportion of participants allocated to the intervention group who complete the intervention, as well as the proportion of missed and incomplete sessions, will be recorded in session logs.
- **Time required for data collection and analysis:** Time for screening, consent, and follow-up will be recorded via timesheets to inform timelines and milestones in a future trial.
- **Feasibility of outcome measures**: The proportion of missing data from each of the outcome measures will be reported to determine the feasibility of collecting complete data using the identified outcome measures.

These measures of feasibility and acceptability will also be explored in more detail in the qualitative process evaluation.

##### Clinical Outcome Measures

Most studies evaluating BA have a follow-up period of less than 3 months (15); therefore, we will administer the clinical outcome measures at baseline and at 3 months post-randomisation. The clinical outcomes will include depression, as measured by the PHQ-9, a commonly used measure of depression that has been psychometrically validated within people who have kidney failure (37). We will also capture anxiety, measured using the GAD-7 (38), health-related quality of life, measured by the KDQOL-SF36 (39), mental well-being, measured by the WEMWBS (40), and adherence to haemodialysis, measured by dialysis attendance captured in the healthcare resource use logs, and completion of full-length sessions. An overview of the schedule of clinical outcome measures is shown in the SPIRIT Figure in Figure 3 (28).

**Figure 3.**
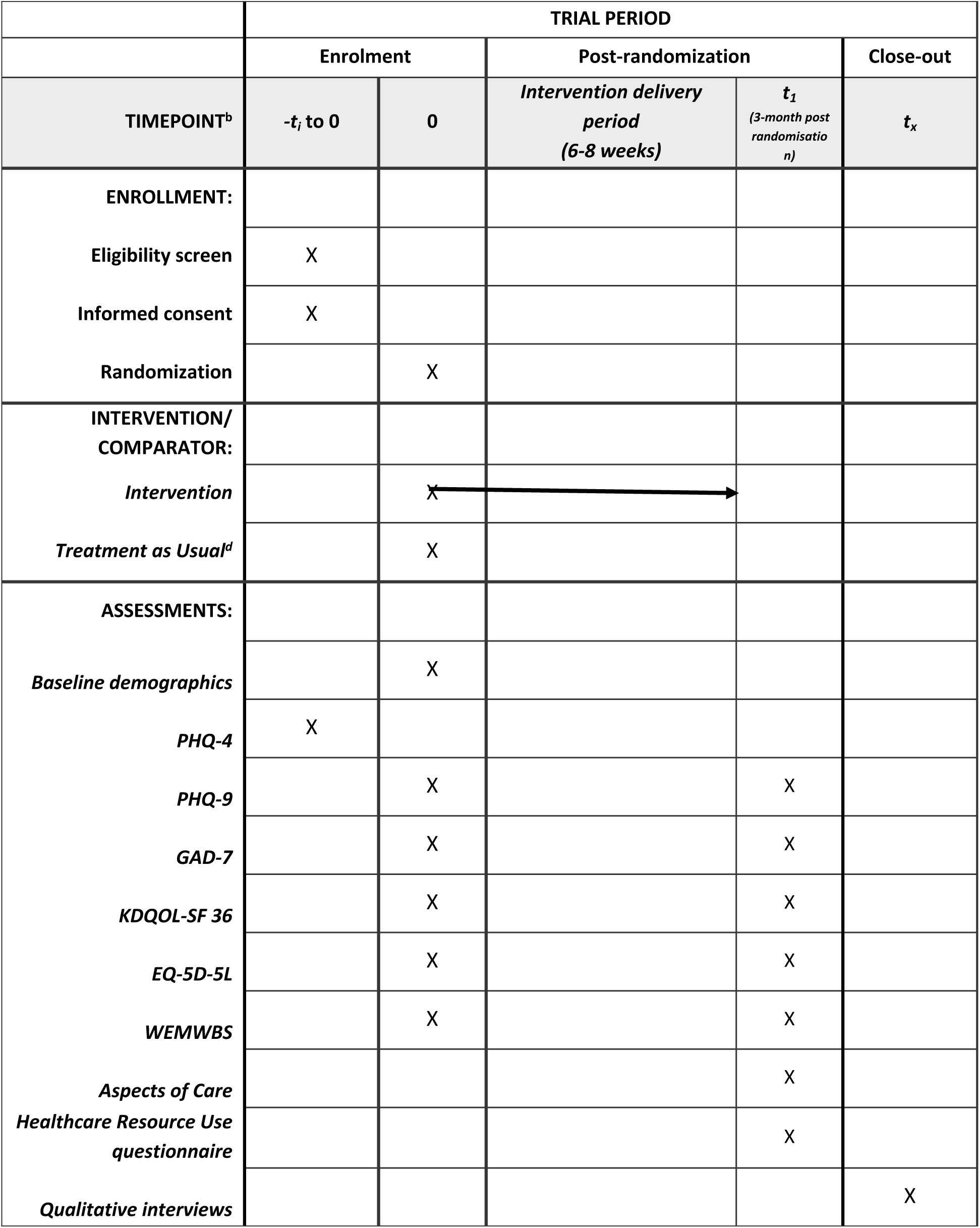
Participant timeline: Schedule of enrolment, interventions, and assessments.

##### Data Analysis

To address the primary feasibility outcomes, a variety of approaches will be used. Descriptive statistics will be used to present baseline demographic and clinical data. Categorical data will be presented as frequencies, and continuous data as means. Summary statistics will be used to describe the proportions of patients approached who consent to screening, are eligible, consent to the study, are randomised, and are retained and followed up. The proportion of people who complete the intervention, the number of intervention sessions received, and the number of adverse events will be summarised. For continuous secondary outcome variables measured at 3-months, effects sizes with 95% CIs will be estimated using linear regression models, with group allocation and the baseline value of the outcome included as covariates.

### Feasibility Economic Evaluation

A feasibility economic evaluation will be embedded within the feasibility cRCT; therefore, participants will provide consent to take part in this element of the study during recruitment to the feasibility cRCT. As this is a feasibility economic evaluation, we will not provide cost-benefit, effectiveness, or utility estimates. Instead, this part of the study aims to explore the feasibility of data collection for an economic evaluation within a definitive trial.

#### Data collection

We will evaluate the feasibility of collecting data related to health and social resource use. Health and social care resource use will be captured using a modified Service Use Questionnaire adapted from previous Service Use logs administered to this population (34, 41). Participants will be asked to complete a retrospective questionnaire to report their healthcare experiences over the past month at baseline and at the 3-month follow-up (41). This will capture information on care received through primary care, outpatient departments, emergency departments, inpatient hospitalisations, and mental health services. The EQ-5D-5L will be administered to participants at baseline and at three-month follow-up, alongside the clinical outcome questionnaires (42).

#### Data Analysis

Descriptive statistics of the response rates, missing data, and costs will be provided (43). This descriptive approach is recommended for feasibility studies, as it provides an overview of the feasibility of the data collection methods without attributing causality to any observed differences in the data (43). Costs will be estimated according to NHS reference costs and presented as a descriptive cost-consequence analysis. Missing data for the service use questionnaires and the EQ-5D-5L will be presented as frequencies to assess the feasibility of data collection in a future cRCT. The acceptability of using service-use questionnaires to collect healthcare-use data will also be explored in the qualitative process evaluation.

### Process evaluation

#### Participants

Participants will include patients who have participated in the feasibility cRCT, their caregivers, and healthcare workers with experience of the BA intervention. We aim to recruit 15 patients who participated in the study, 10 healthcare workers who either delivered the intervention or had the opportunity to work alongside its delivery, and five carers of individuals who received the integrated BA intervention (44).

Healthcare workers will be eligible for participation in the study if:

- They are a member of a multidisciplinary team, including nurses, medical doctors, healthcare support workers, physiotherapists, and social workers.
- They have had exposure to the intervention, either as a facilitator who delivered it or as a team member present in the unit during its delivery.
- They have at least three months of experience within the kidney care setting.

Carers will be eligible for participation in the study if:

- They are a family member or friend who provides support to a participant in the study who received the integrated BA intervention.
- They are over 18.
- They have the capacity to provide informed consent.

We will recruit patients from both the intervention and control groups to assess the acceptability of the intervention and trial procedures.

#### Recruitment

Patients recruited into the feasibility cRCT will have the opportunity to participate in the qualitative process evaluation. They will be provided with a further participant information sheet providing an overview of the qualitative process evaluation, which will have been developed in collaboration with our PPI panel. Consent will be obtained before the interview begins. We will attempt to recruit a diverse range of patient participants at the two sites to ensure that the interviews capture a broad spectrum of experiences across demographic groups.

Healthcare workers who have delivered the BA intervention or were on shift when it was delivered will be approached directly by a researcher and invited to participate in the process evaluation. They will receive a participant information sheet and will be given at least 24 hours to decide whether they wish to participate.

Carers will be recruited through patients who received the BA intervention as part of the feasibility cRCT. Participants who received the intervention will be provided with a recruitment pack to provide to a family member or friend. All participant-facing materials will be developed in collaboration with our PPI panel.

#### Data collection

We aim to conduct 35 semi-structured interviews, comprising 15 with participants who have taken part in the study, 15 with healthcare workers with experience of the intervention (including intervention facilitators), and 5 with carers of participants who received the intervention. Semi-structured interviews will enable an in-depth exploration of experiences within the feasibility cRCT and allow data collection to align with healthcare workers’ work commitments and patients’ treatment burden. Interviews will be conducted in a suitable location for participants. In previous feasibility studies conducted in haemodialysis units, this is frequently during the workday or during haemodialysis treatment (45).

The semi-structured interviews will follow a topic guide structured around the theoretical framework of acceptability (46). They will focus on the acceptability of the integrated BA intervention and the trial procedures (including screening with the PHQ-4). These interview guides will be informed, developed, and refined through consultation with the PPI panel. The interviews will be audio-recorded and transcribed verbatim.

#### Data Analysis

Interviews will be recorded verbatim using encrypted recording devices. Thematic analysis will be conducted, using NVivo, to identify overarching themes in the data. The interviews will be coded line by line, and these codes will then be reviewed to group relevant codes. These groups will then be iteratively revised to identify the common themes and sub-themes in the data (47). We will present the themes to our PPI panel to explore their perspectives on the findings, how they can inform a definitive trial, and their relevance to practice.

### Progression Criteria

Progression to a definitive cRCT will be determined by recruitment rates and the acceptability of the methods and intervention for patients and staff. Both quantitative and qualitative data will be used to determine the feasibility of a cRCT (34, 48):

- Recruitment

◦ 75-100% of the target sample size recruited across the two sites will result in progression to a definitive trial.
◦ 50-74% of the target sample size recruited across the two sites will result in progression to a full trial, following appropriate amendments to address barriers to recruitment.
◦ 25-50% of the target sample size recruited across the two sites will result in progression to a pilot study following review by the study steering committee and future co-applicants to ensure that adequate revisions are made to address barriers to recruitment.
◦ <25% of the target sample size recruited will result in the study not progressing to a full trial.
- Randomisation

◦ Evidence of substantial or consistent contamination from the method of clustering (either due to high rates of contamination or evidence of significant contamination from qualitative interviews) will result in a review of the randomisation and clustering method, before progression to a definitive trial.
- Intervention

◦ Progression to a full trial will be contingent on the acceptability of the integrated BA intervention for both patients and healthcare professionals. This will be determined by exploration within the qualitative process evaluation. Any necessary modifications or adaptations will be made in collaboration with our PPI panel.
- Data collection

◦ The use of the clinical outcome measures in a future trial will depend on their acceptability, determined by the proportion of missing information, and through the qualitative process evaluation.

## Discussion

People with kidney failure receiving haemodialysis are at higher risk of developing mental health conditions, such as depression(3); despite this, there is currently limited access to support and specialist mental health services. Alongside current barriers to accessing support, such as long waiting times, patients spend a significant portion of their week attending dialysis appointments(6, 7). One potential solution is to explore ways to improve access to mental health support and treatment within their existing treatment regimen.

While BA is a NICE-recommended treatment for depression in adults, including those with co-occurring long-term conditions(17), it has never been delivered or evaluated for patients with kidney failure during their routine haemodialysis appointments(10, 49). Therefore, our primary aim isn’t to explore its effectiveness, but rather to first assess the feasibility and acceptability of delivering BA during haemodialysis. We will explore this through various methods; firstly, a pilot cRCT to determine the feasibility of conducting a definitive cRCT to evaluate the BA intervention, secondly, a qualitative process evaluation to explore the acceptability of the intervention and methods, and finally, a feasibility economic evaluation to explore the best methods for evaluating cost-utility in a definitive trial.

A key strength of this study is that the intervention was adapted in close collaboration with patients and carers, with the aim of further refinement following delivery within the pilot cRCT. The protocol follows the Medical Research Council’s (MRC) guidance on the evaluation of complex interventions(27), using a comprehensive mixed-methods approach to explore feasibility and acceptability. While the study will involve only two sites in England and we do not have the resources to translate and culturally adapt the intervention, which limits the generalisability of the findings, we have deliberately selected sites that serve different demographics. Therefore, the feasibility study will provide us with an understanding to refine the protocol and intervention for a future definitive cRCT.

## Data Availability

No datasets were generated or analysed during the current study. All relevant data from this study will be made available upon study completion.

## Acknowledgements

This project is funded by the NIHR Research for Patient Benefit (NIHR209283). The views expressed are those of the author(s) and not necessarily those of the NIHR or the Department of Health and Social Care. We would like to thank our Patient and Public Involvement panel for their ongoing support and guidance.

